# Longitudinal changes in retinal microvascular during healthy pregnancy measured by Adaptive Optics imaging – the PAPYRUS study

**DOI:** 10.64898/2025.12.02.25341523

**Authors:** Anne Dathan-Stumpf, Emilie Martini, Holger Stepan, Radim Kolar, Franziska G. Rauscher

## Abstract

**Background:** Adaptive Optics retinal imaging (AO) enables high-resolution visualization of the retinal microvasculature providing insights into systemic vascular health. Currently, no studies exist describing changes in wall-to-lumen ratio (WLR) during pregnancy, neither during the physiological course of pregnancy nor in pregnancy-associated complications.

**Methods and Material:** This single-centre, prospective study at the Leipzig University Hospital, Germany, examines longitudinal changes in retinal microvasculature by employing AO (rtx1e, Imagine Eyes, Orsay, France) to calculate the WLR of an arteriole within few seconds. The study examined a well-phenotyped cohort of 495 primarily Caucasian healthy pregnant women, with 584 measurements taken from the first to the third trimester. The WLR was automatically determined using the nnUNet deep learning model.

**Results:** As demonstrated in the linear mixed-effects model, gestational age exhibited the most significant effect on WLR (p < 0.001). Changes in WLR were also significantly associated with maternal age (p = 0.04) and mean arterial blood pressure (p < 0.001). Primiparous women had significantly lower WLRs at baseline (p < 0.01) and exhibited significantly steeper increases in WLR throughout gestation (p = 0.01) than nulliparous women.

**Conclusion:** We identify WLR as a sensitive marker for imaging microvascular remodeling, serving as an indicator of adaptation to physiological pregnancy. Normal pregnancy is associated with an instant change of the retinal microvasculature indicated by an increase of WLR. Further studies are required to investigate the postpartum course of WLR and establish whether these changes are reversible.

## Introduction

Pregnancy is widely recognized as a “natural stress test” for the maternal cardiovascular system. Hemodynamic, hormonal, and metabolic changes that occur throughout gestation impose a significant burden on vascular homeostasis, requiring adaptive responses to ensure adequate maternal and fetal perfusion ^1^. The vascular remodeling process involves structural and functional changes in both large and small arteries and veins. These changes include increases in luminal circumference and length, tissue and cellular hypertrophy, endothelial and vascular smooth muscle hyperplasia, and matrix remodeling ^2–4^, with remodeling in large vessels being time-consuming to measure by various methods such as arterial tonometry or carotid intima-media thickness measurement ^5^.

The eye is increasingly recognized as a non-invasive window to the microvasculature, offering insights into systemic health conditions ^6–8^. This is primarily due to the ability to directly visualize and quantify the microvasculature of the retina using techniques such as Adaptive Optics (AO) imaging, which provides high-resolution, non-invasive and in-vivo images of microscopic structures in the human retina ^9,10^. The ability to identify subtle structural changes before clinical symptoms appear enables earlier intervention and better monitoring of cardiovascular risk ^11^. AO imaging has been instrumental in advancing the understanding of various conditions, including diabetes, stroke, and dementia, by allowing to observe changes in the retinal microvasculature ^10,12,11^. Therefore, AO measurements could serve as a noninvasive biomarker for maternal microvascular health and pregnancy outcome prediction ^12^. In addition, the technique is suitable to monitor disease progression and response to treatment without discomfort to the patient ^13^. In a preliminary study by our group, demonstrated relevant longitudinal changes in the retinal microvasculature using AO in a healthy pregnant woman with a twin pregnancy ^14^. In contrast to other established ophthalmic methods, AO allows microscopic resolution of a single arteriole with visualization of the vessel wall and lumen.

Besides this single case analysis, to our knowledge, there is no other publication presenting (longitudinally) investigated changes in retinal microvasculature during pregnancy measured by AO.

The aim of this prospective longitudinal study was to map changes in retinal microvasculature, as an expression of systemic vascular remodeling, across trimesters in the physiological course of pregnancy. The definition of such normative data on microvascular changes in healthy pregnant women will allow the description of changes in pregnancy-associated complications such as hypertensive pregnancy disorders or gestational diabetes mellitus. In addition, these data could be used as a basis for postpartum follow-up measurements to predict individual cardiovascular risk after pregnancy-associated complications.

## Methods

### Study Population

This monocentric, prospective study analyzed the longitudinal changes in retinal microvasculature during pregnancy exclusively in healthy pregnant women. Participation in the study was possible from the first trimester, and follow-up measurements were performed longitudinally at least once in each subsequent trimester. The first trimester was defined as gestational age up to 13.6 weeks of gestation (wog), the second trimester between 14.0 and 27.6 wog and the third trimester from 28.0 to 42.0 wog.

Pregnant women were defined as healthy if they did not have a hypertensive pregnancy disorder (pre-eclampsia, pregnancy-induced hypertension, chronic hypertension), HELLP-syndrome, intrauterine growth restriction, pre-existing diabetic metabolic disease, gestational diabetes (GDM) or cholestasis of pregnancy. Women who had a hypertensive pregnancy disorder or gestational diabetes in previous pregnancies were also excluded from this analysis. Women who were enrolled early in pregnancy as healthy pregnant women and were subsequently diagnosed with a pregnancy-related complication named above were excluded for this analysis. Additionally, we excluded patients with pre-existing (surgically corrected) structural heart diseases or heart failure (according to NYHA criteria), pre-existing nephropathies/glomerulonephritis, known vasculitides and/or collagenoses (e.g. systemic lupus erythematosus), and use of any type of lipid-lowering agents and diuretics. These exclusion criteria were chosen because these conditions lead to a pregnancy-independent organ damage and an increased risk of cardiovascular diseases. We also excluded fetal chromosomal abnormalities or genetic defects associated with fetal growth restriction, and patients with suspected major fetal structural defects (e.g., heart failure), which can lead to fetal ascites and hydrops fetalis and are therefore associated with maternal mirror syndrome. Furthermore, as changes in retinal microvasculature are described for neuro- and musculodegenerative diseases, women with such diseases were also excluded from the analysis ^15,16^. In addition, women with pronounced hyperopia or myopia more than 6 dioptres, laser refractive surgery treatment, or eye disease (e.g. retinal degenerative changes, glaucoma) were excluded.

Additionally, a control group of non-pregnant, eye healthy, non-hypertensive and metabolically healthy Caucasian women was recruited. These women received their AO measurement at a single visit.

### Investigation parameters

All maternal and pregnancy-related data were collected prospectively. During the study, participants were phenotyped using a very detailed family and medical history, including information on previous pregnancies, and the current pregnancy, current medication history, smoking status, gestational age and week of pregnancy, results of the 50g screening test for GDM and, if abnormal, the results of the 75g oral glucose tolerance test ^17^, as well as body mass index before pregnancy, at the time of measurement and at delivery. In addition, all patients had two standardized blood pressure measurements taken at each visit after a five-minute rest ^18^, with the first measurement discarded and only the second measurement included in the analysis ^19–21^.

During the first to third trimester visits, pregnant women also underwent standardized measurements of both uterine arteries with documentation of the pulsatility index (PI) ^22^. The PI of the uterine artery is a validated and reliable tool for detecting increased impedance to blood flow, which is associated with increased placental resistance ^23^.

Recruitment took place from April 2023 to May 2025 through the outpatient clinic of the Leipzig University Hospital, Department of Obstetrics, Germany. Women could be enrolled at any stage of pregnancy; singleton and multiple pregnancies were evaluated separately.

### Eye measurements by Adaptive Optics retinal imaging (rtx1e) and Wall-to-Lumen ratio (WLR)

The Adaptive Optics fundus photography device (rtx1e, Imagine Eyes, Orsay, France) is a retinal camera at high-resolution producing en-face images of the retina (4°x4°), facilitating non-invasive and in-vivo examination of microscopic walls of retinal arterioles without the use of contrast agents ^24,25,12,26–28^. AO retinal imaging can achieve optical resolutions of 1-2 μm in the living human eye ^9^ by correcting optical aberrations, allowing for the analysis of vascular wall morphology and other parameters ^29^. The method can be used to reliably determine inner and outer vessel diameters, the vessel wall thickness, the Wall-Cross-Section-Area and the Wall-to-Lumen Ratio (WLR) ^30^. The device’s software enables repeated imaging of the same retinal region, which is crucial for longitudinal measurements in pregnancy and allows monitoring of disease progression and response to treatment ^29^. Furthermore, WLR assessments of the obtained images are accurate and can be reproduced reliably ^31^, with low variability between observers. This enables differentiation between hypertrophic and eutrophic remodeling processes ^32,12^. Meixner et al. reported an intra-observer variability ∼6% for wall thickness and ∼2% for total of wall and lumen measurements ^12^. In addition, AO retinal imaging is fast and non-invasive and thus well tolerated by patients ^9^. Acquisition of a vascular image of the retinal microvasculature using AO takes 5 seconds, instrument setup and focusing prior to that is also quickly accomplished. As studies have shown that changes during pregnancy affect both eyes simultaneously ^26^, only the right eye of all participants was measured in the current study.

In this study, we employed an automatic segmentation approach using the nnUNet ^33^ deep learning model ^34^ to determine the WLR values in retinal imaging. The nnUNet based model was trained on manually segmented arteries and walls in AO images, and it accurately identified areas of the vessel walls and lumen. From these segmented areas, the WLR was calculated by automatic measuring the thickness of the arterial wall and the diameter of the vessel lumen. For arteries with visible walls on both sides, the WLR was determined using the combined thickness of both walls relative to the lumen diameter. In cases where only one side of the arterial wall was clearly visible, the WLR was adjusted accordingly (Figure 1 A and B). This automated method provides a reliable and efficient approach for clinical assessment of retinal microvascular structures ^34^.

**Figure 1.**
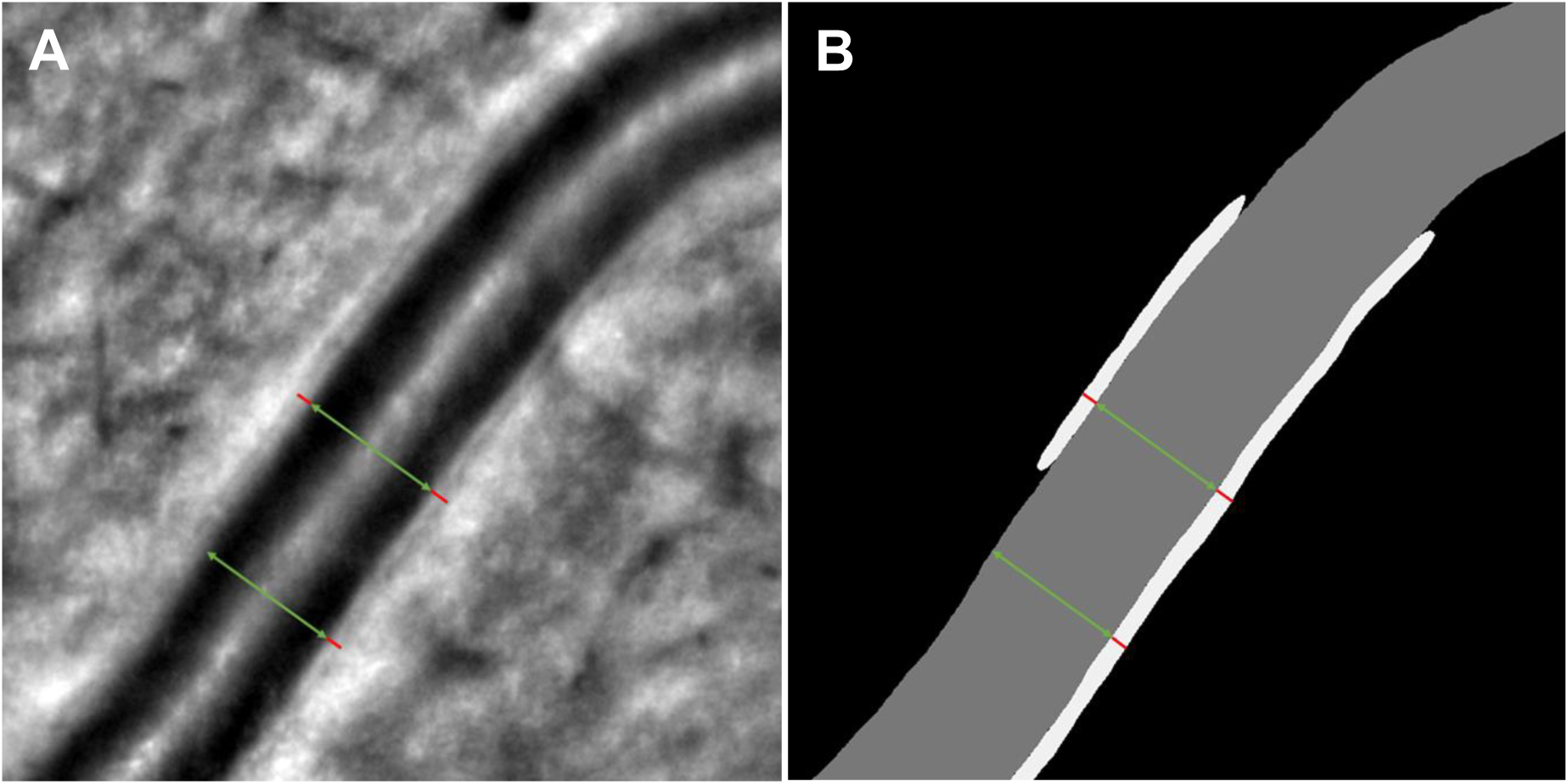
A) Original image of a retinal arteriole: The green arrow represents the lumen and red lines represent the wall of the vessel ^14^. B) Automatic segmentation result of this arteriole: An automatic measurement of lumen and wall is applied on several place along the segmented area. Prior to grading, images with poor quality were removed manually (N = 53).

#### Statistical analysis

The programming platform Matlab R2024a was utilized for the analysis and graphical representation of the data. Standard statistical methods with a significance level of 5% (p = 0.05) were employed for all tests. Linear mixed-effects models were employed to analyze the effects of various predictor variables. In the linear mixed-effects model WLR was designated as the response variable, the potential influencing variables such as gestational age serving as predictor variables. Furthermore, linear regression analyses were conducted. The likelihood ratio test and the partial F-test were utilized to compare the full and reduced models in logistic and linear regression, respectively. Pearson’s correlation coefficient was utilized to depict linear relationships between dichotomous variables scaled to the interval level.

#### Ethics

The measurements were performed as part of the PAPYRUS study, which aims to predict the individual cardiovascular risk after hypertensive pregnancy disorders and GDM by longitudinal changes in the retinal microvasculature (German Clinical Trial Register number: DRKS00032530). Written informed consent was obtained from all participants. Research related to human use has been complied with all the relevant national regulations, institutional policies and in accordance the tenets of the Helsinki Declaration, and has been approved by Leipzig University’s institutional review board (IRB00001750, 052/23-ek).

## Results

### Baseline characteristics of the current study population

A total of 495 pregnant women, as well as 76 non-pregnant women, all were ophthalmologically and systemically healthy, were recruited and examined between April 2023 and May 2025. Baseline characteristics of the final cohort as well as stratified by trimester of well phenotyped healthy pregnant women are presented in Table 1. The majority of participants were women of Caucasian origin (90%). Almost half of the participants had never given birth, and just under 40% had given birth to one child. The mean PI values of the uterine arteries, which are contingent on gestational age, corresponded to the reference values for healthy pregnant women of Caucasian origin ^22^.

**Table 1.** This description provides an overview of the cohort of healthy pregnant women according to certain characteristics, with the data divided by trimester of pregnancy. Some patients only entered the study in the third trimester, thus, measurements in the first and second trimesters were skipped. For N=134 healthy pregnancies, the mode of delivery was not documented due to the patients either giving birth in other hospitals or being still pregnant at the time of analysis, with study recruitment ongoing.

|  | Parameters | Description |
| --- | --- | --- |
| **Total study population** | Number of subjects | 495 |
|  | Number of measurements | 584 |
|  | Maternal age [years] (mean±SD) | 32.1 ± 5.5 |
|  | Ethnicity [N (%)] |  |
|  | European/ Caucasian | 444 (89.7) |
|  | Asian | 18 (3.6) |
|  | Black | 10 (2.0) |
|  | Arabs | 16 (3.3) |
|  | Mixed | 7 (1.4) |
|  | Gravidity [N (%)] |  |
|  | 1 | 183 (37.0) |
|  | 2 | 167 (33.7) |
|  | 3 | 89 (18.0) |
|  | ≥ 4 | 56 (11.3) |
|  | Parity [N (%)] |  |
|  | 0 | 237 (47.9) |
|  | 1 | 190 (38.4) |
|  | ≥ 2 | 68 (13.7) |
|  | BMI before pregnancy [kg/m<sup>2</sup>] (mean±SD) | 24.4 ± 4.9 |
|  | BMI at delivery [kg/m<sup>2</sup>] (mean±SD) | 29.5 ± 4.9 |
|  | Singleton pregnancy [N (%)] | 466 (94.1) |
|  | Twin pregnancy [N (%)] | 29 (5.9) |
|  | Value socio-economic status | 15.7 ± 3.4 |
|  | gestational age at delivery [weeks.days] | 39.2 |
|  | vaginal birth [N (%)] | 232 (64.3) |
|  | vaginal operative [N (%)] | 34 (9.4) |
|  | cesarean section [N (%)] | 95 (26.3) |
|  | birth weight [g] | 3387 ± 512 |
| **1st trimester** | Number of subjects / measurements [N] | 45 / 46 |
|  | gestational age [weeks.days] | 12.6 |
|  | systolic blood pressure [mmHg] (mean±SD) | 116.4 ± 12.3 |
|  | diastolic blood pressure [mmHg] (mean±SD) | 72.9 ± 9.1 |
|  | MAP [mmHg] | 87.4 ± 10.1 |
|  | BMI [kg/m<sup>2</sup>] (mean±SD) | 24.8 ± 5.2 |
|  | PI Aa. uterinae (mean±SD) | 1.18 ± 0.41 |
| **2nd trimester** | Number of subjects / measurements [N] | 173 / 175 |
|  | gestational age [weeks.days] | 21.3 |
|  | systolic blood pressure [mmHg] (mean±SD) | 114.8 ± 11.2 |
|  | diastolic blood pressure [mmHg] (mean±SD) | 69.9 ± 8.3 |
|  | MAP [mmHg] | 84.9 ± 10.1 |
|  | BMI [kg/m<sup>2</sup>] (mean±SD) | 26.7 ± 6.0 |
|  | PI Aa. uterinae (mean±SD) | 0.91 ± 0.27 |
| **3rd trimester** | Number of subjects / measurements [N] | 360 / 363 |
|  | gestational age [weeks.days] | 34.6 |
|  | systolic blood pressure [mmHg] (mean±SD) | 116.5 ± 11.1 |
|  | diastolic blood pressure [mmHg] (mean±SD) | 73.2 ± 8.5 |
|  | MAP [mmHg] | 87.6 ± 9.9 |
|  | BMI [kg/m<sup>2</sup>] (mean±SD) | 28.7 ± 4.9 |
|  | PI Aa. uterinae (mean±SD) | 0.76 ± 0.27 |

N – number; g – gram; SD – standard deviation

#### Effect of gestational age on changes of the WLR

The gestational age has a significant effect (p <0.001) on the WLR. We analyzed the effect of gestation week on WLR during pregnancy using a linear mixed-effects model.

For *WLR = Baseline + Slope × Week of gestation* we found that WLR = 0.1858 + 0.000445 × *Week of gestation* (Table 2, Figure 2).

**Figure 2.**
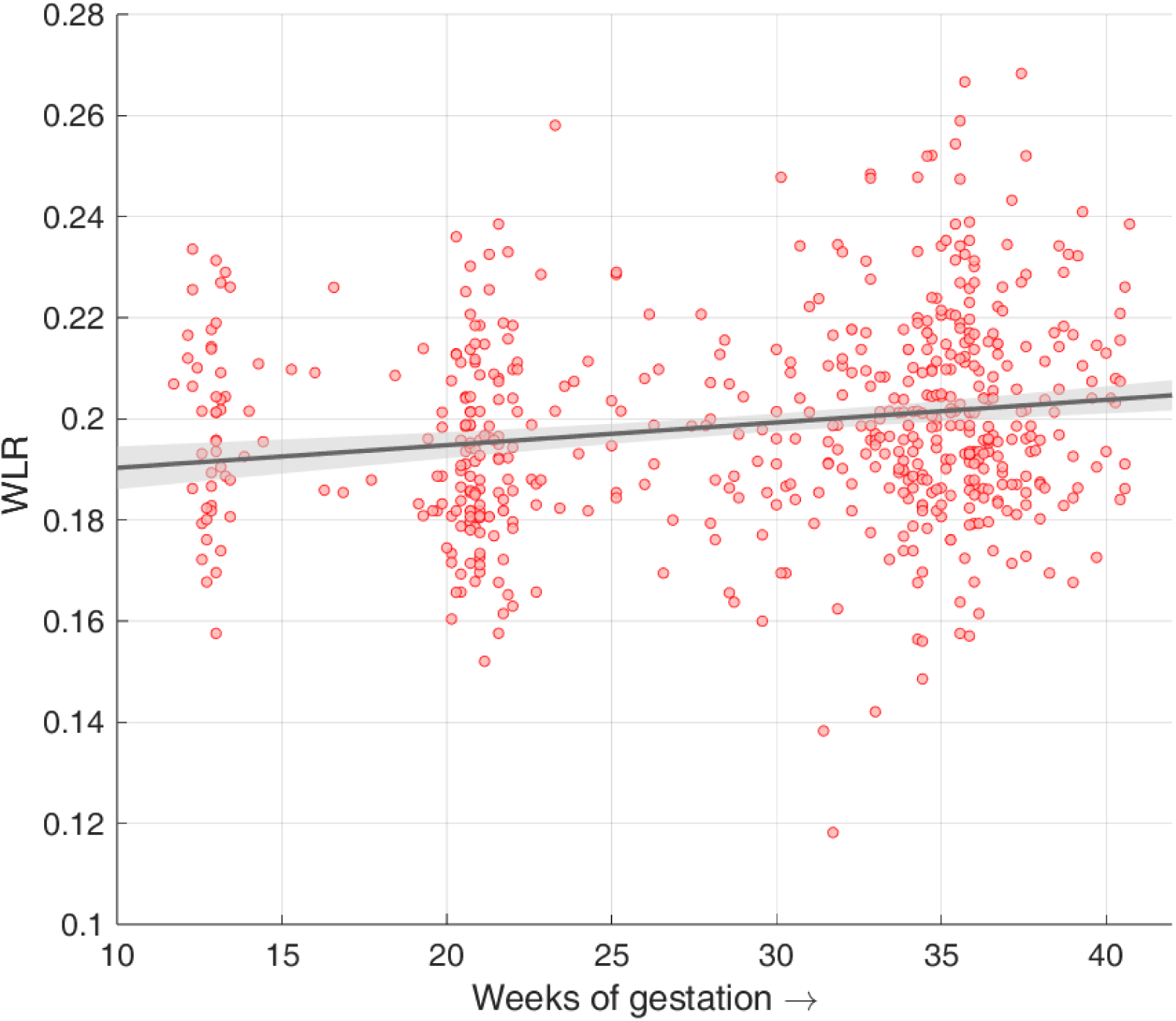
Changes of WLR during pregnancy: The scatterplot of the WLR in the full healthy pregnancy cohort (N = 495) over gestational age until 42 weeks of gestation demonstrates a significant increase in WLR. The red points demarcate the individual WLR measurements (N = 584). The black line signifies the linear regression based on the data sample, whilst the grey area denotes the 95% confidence interval of the linear fit based on bootstrapping (see Figure S4 for stratification in groups of parity).

**Table 2.**
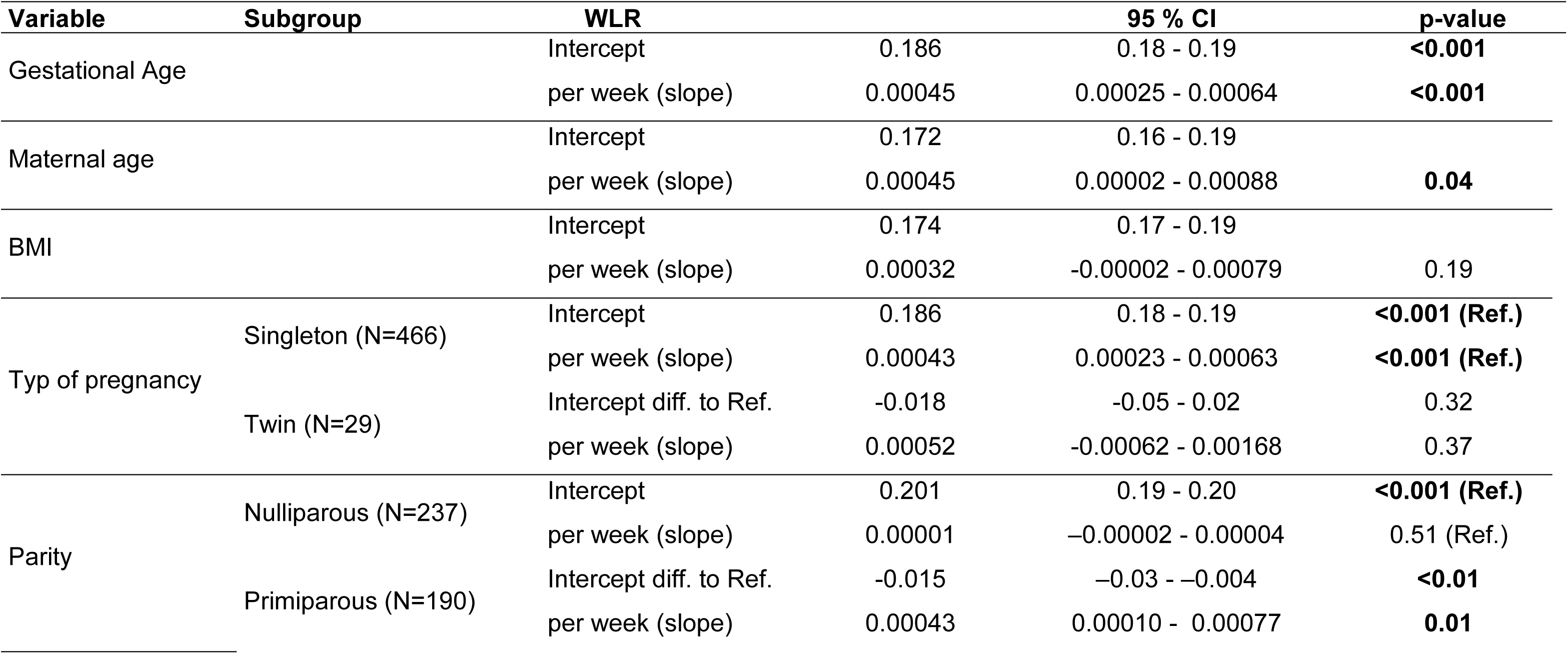
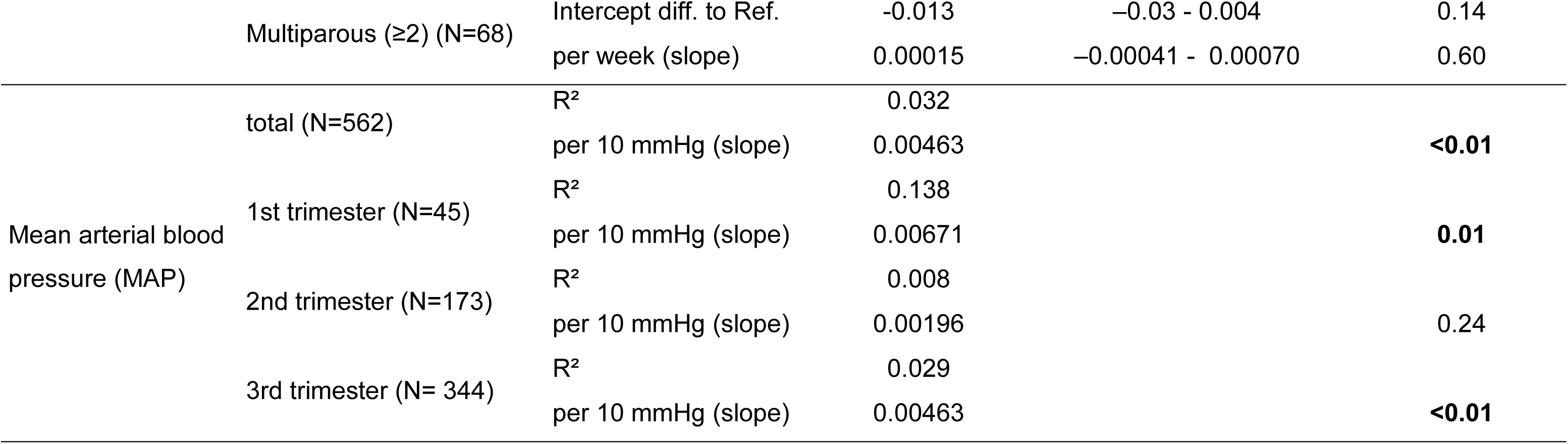
Summary of the analyses of the potential influencing factors on the longitudinal development of WLR throughout pregnancy. The intercept represents the estimated WLR at conception (0 weeks of gestation), the intercept difference to reference (diff. to ref.) points out the difference between the WLR to the respective reference cohort (Ref.). In cases where the “Intercept diff. to Ref.” is given, both the 95% CI and the p-value also refer to this difference between the two compared groups. The slope represents the estimated increase in WLR per week of gestation within a linear mixed-effects model. Significant values are highlighted in bold.

Using the linear regression model to estimate the WLR in week 0 of gestation and setting 0.1858 as the baseline (corresponding to 100%), the increase in WLR at the end of pregnancy for mean gestational age of delivery (39.2 wog) is 0.2033, which corresponds to a 9.4% change.

The mean values of the WLR, depending on the selected gestational age intervals, are shown in Table 3, when compared to the healthy, non-pregnant reference group. A comparison of the plotted WLR over the course of pregnancy was made with that of the non-pregnant reference group. A statistically significant difference was found between the two groups from 32th until the 42th week of gestation.

**Table 3.** The following table illustrates the mean WLR values as a function of gestational age (Figure 3 for graphical representation). The mean WLR of the healthy, non-pregnant control cohort is utilized as a reference. From the 32th until the 42th week of gestation the WLR in healthy pregnant women differed statistically significantly from the control cohort of non-pregnant women.

| **Gestational age** | **N measurements** | **Mean** | **Min** | **Max** | **CI lower** | **CI upper** |
| --- | --- | --- | --- | --- | --- | --- |
| 11.0-13.6 | 46 | 0.198 | 0.158 | 0.234 | 0.193 | 0.204 |
| 14.0-21.6 | 131 | 0.193 | 0.152 | 0.239 | 0.190 | 0.196 |
| 22.0-27.6 | 44 | 0.199 | 0.163 | 0.258 | 0.193 | 0.204 |
| 28.0-31.6 | 53 | 0.193 | 0.118 | 0.248 | 0.187 | 0.199 |
| 32.0-33.6 | 53 | 0.201 | 0.142 | 0.248 | 0.196 | 0.206 |
| 34.0-35.6 | 128 | 0.203 | 0.149 | 0.267 | 0.200 | 0.208 |
| 36.0-37.6 | 84 | 0.202 | 0.161 | 0.333 | 0.197 | 0.207 |
| 38.0-39.6 | 32 | 0.205 | 0.168 | 0.282 | 0.196 | 0.214 |
| 40.0-42.0 | 13 | 0.207 | 0.184 | 0.239 | 0.197 | 0.217 |
| Control | 76 | 0.194 | 0.158 | 0.235 | 0.190 | 0.197 |

N measurements – number of measurements; Min – minimum; Max – maximum; CI – confidence interval (lower at 2.5% and upper at 97.5%)

For the longitudinal study, a subset of women was selected who were examined at least twice, with at least 16 weeks between the first and last examinations. This was considered to be slightly more than one trimester. A total of 14 cases were selected for the purpose of studying the change in WLR value on an individual basis. These values are plotted in Figure S1. We also computed the relative WLR change, or WLR gain, as follows:

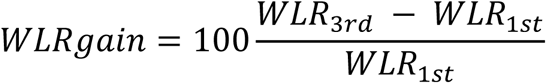

A positive WLR change was confirmed in all cases, with a mean WLR gain of 8.0 % (Table S1).

### Effect of maternal age and BMI on WLR

To evaluate the potential influence of age or BMI before pregnancy on WLR, we performed separate linear regression analyses. A small but statistically significant positive association was observed between age and WLR (0.00045, p = 0.042), (Table 2) suggesting that older maternal age may be linked to a slightly higher WLR. However, no association was observed for BMI before pregnancy and WLR (0.000316, p = 0.19) (Table 2). These findings were consistent with the visual inspection of scatter plots with linear fits (Figure S2).

Given the unavailability of a WLR baseline value (measured at conception in week 0), it should be noted that for this analysis, the WLR value measured in the third trimester was employed utilizing the large number of samples collected during the last trimester. While a small correlation between age and WLR was identified, the development of a correction model necessitates the availability of baseline values of WLR at conception or before pregnancy. In addition, given the relatively limited age range (predominantly between 20 and 40 years), it was deemed unnecessary to correct WLR for age, in our study for subsequent analyses.

We investigated whether including BMI improves the prediction of the weekly WLR change during pregnancy. A linear mixed-effects model incorporating gestational week and random subject intercepts was expanded to include BMI as a fixed effect. However, the fixed effect of BMI was not statistically significant (p = 0.089). Model comparison indicated only a marginal improvement in fit when BMI was added (ΔAIC = 0.5; likelihood ratio test: χ²(1) = 2.50, p = 0.114). Furthermore, the interaction between BMI and gestational week was not significant (χ²(1) = 0.44, p = 0.51), suggesting that maternal BMI does not affect the longitudinal trajectory of WLR during pregnancy.

### Comparative analysis of singleton versus twin pregnancies

We analyzed the increase of WLR across gestation in single (N = 466) and twin pregnancies (N = 29) using linear mixed-effects modeling. In single pregnancies, WLR demonstrated a statistically significant increase with increasing gestational weeks. When comparing singleton and twin pregnancies, neither their baseline WLR (corresponding to estimate at conception; 0.186 vs. 0.168) nor the rate of increase of WLR over time (0.00043 vs. 0.00052) differed significantly between groups (Table 2). As can be seen in Figure S3, the interaction term between gestational week and pregnancy type was not significant, indicating that the temporal trajectory of WLR is similar for both single and twin pregnancies. This supports the conclusion that WLR evolves similarly in either group over time (Figure S3).

### Influence of parity on the WLR

We used a linear mixed-effects model to evaluate whether parity influenced WLR trajectories across gestation, taking into account gestational week, parity (nulliparous, primiparous, multiparous), their interaction, and random subject intercepts. The mean maternal age (± SD) in the respective parity groups was comparable (nulliparous 31.2 ± 5.4, primiparous 33.2 ± 4.9 and multiparous 33.8 ± 5.2). Compared with nulliparous women (as reference group), those with one previous pregnancy had a significantly lower WLR at baseline (0.186, p <0.01). However, the latter exhibited a significantly steeper increase in WLR throughout gestation (0.00043, p = 0.01). Women with two or more previous pregnancies also showed the lower WLR values at baseline (0.188) described in primiparous women, but increased much less steeply in the further course of the pregnancy (Table 2, Figure S4).

#### Correlation with mean arterial blood pressure (MAP)

Pearson correlation analyses were performed to analyze the relationship between mean arterial pressure (MAP) and WLR. A positive trend between MAP and WLR (R² = 0.032; p < 0.01) was observed for the entire cohort, regardless of gestational age. When gestational age was taken into account, however, this correlation was strongest in the first trimester (R² = 0.131, p= 0.01) and the third trimester (R² = 0.029, p= 0.03) (Table 2, Figure 3; Figure S5). As can be seen from Figure S6 A-C, when parity was taken into account, Pearson correlation analyses revealed a positive correlation between MAP and WLR only among nulliparous women (R^2^ = 0.66; p = 0.008), with a similar but not statistically significant trend was observed among primiparous (R^2^ = 0.06; p = 0.54) or multiparous (R^2^ = 0.13; p = 0.34) women.

**Figure 3.**
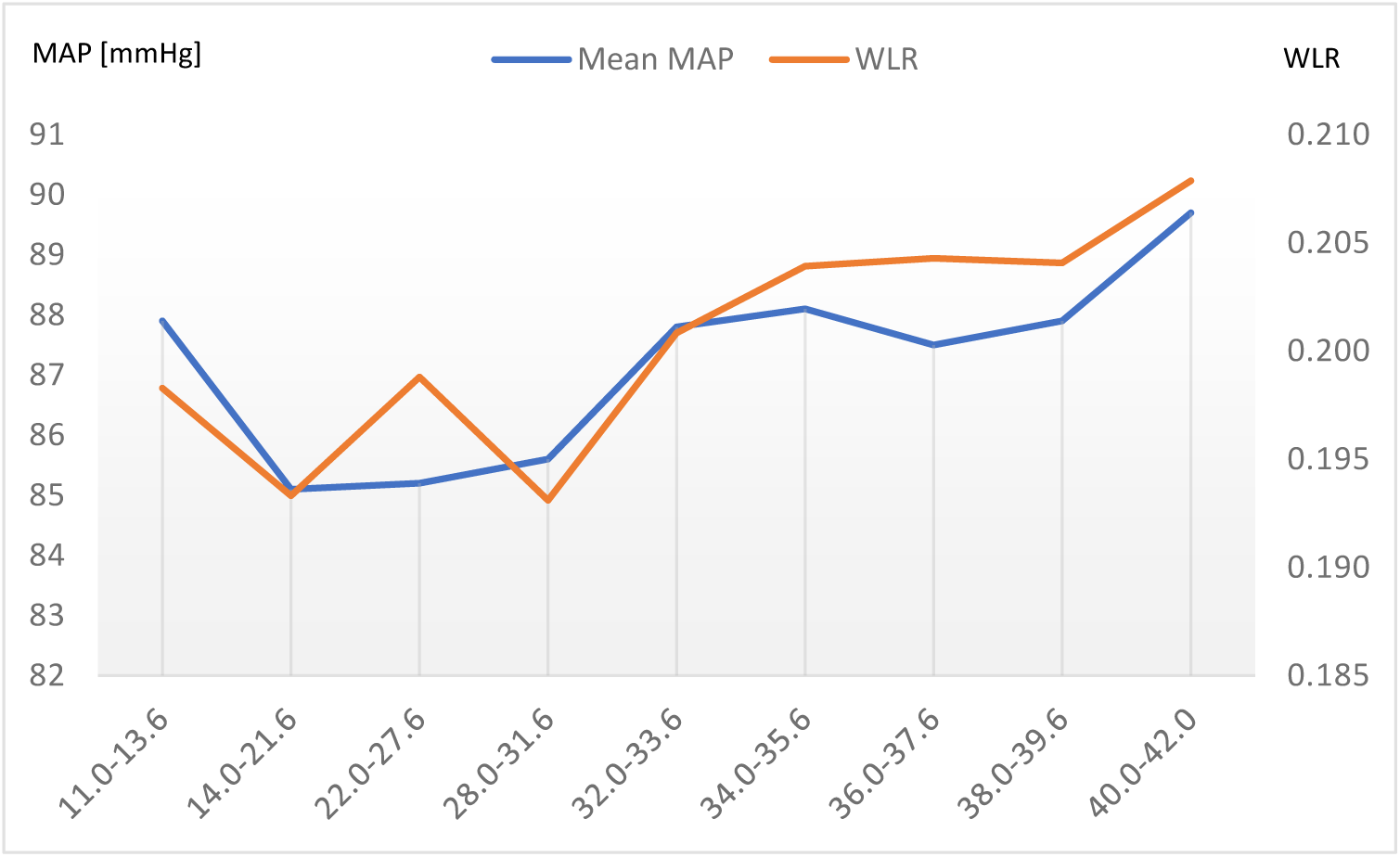
Relationship between WLR and mean arterial blood pressure (MAP): A significant correlation (R²= 0.032; p < 0.01) was found between MAP and WLR in the first and third trimesters (trimester analyses see Figure S5). This increase in WLR with gestational age indicates cardiovascular remodelling in response to pregnancy. Note that WLR generally increases with weeks of gestation (Figure 2). The relationship of MAP and WLR depicted in Figure 3 is averaged across all healthy pregnant women investigated. When stratified by parity (see Figure S6) the dip in WLR in the first weeks of gestation was driven by multiparous women.

### Correlation with pulsatility index of Aa.uterinae

Linear regression analysis revealed a significant negative correlation between gestational age and the uterine artery pulsatility index (−0.012, p < 0.001). The model explained approximately 10% of the variance in PI values (R² = 0.107). As gestational age increased, PI values decreased, which is consistent with the expected physiological changes that occur during physiological pregnancy. The relationship between averaged PI and WLR at the time of measurement, depending on gestational age, is shown in Figure S7.

## Discussion

To the best of our knowledge, first, there are no studies which demonstrate changes in retinal microvasculature during pregnancy, and, second, outside of our own work no studies obtain in-vivo high-resolution vascular data measured by AO with cellular-level resolution. In this prospective and longitudinal study, we demonstrate significant changes in WLR in a well-phenotyped cohort of healthy pregnant subjects.

The predictive capacity of WLR for cardiovascular outcomes stems from its pathophysiological linkage with systemic microvascular remodeling. An elevated WLR reflects increased peripheral vascular resistance, a key determinant of afterload and long-term target organ damage. Importantly, retinal arteriolar changes mirror analogous alterations in other vascular beds, such as coronary and renal microcirculation, making retinal WLR a surrogate marker of generalized microvascular health. Longitudinal studies have demonstrated that higher retinal WLR values are associated with increased risk of stroke, myocardial infarction, and progression of chronic kidney disease ^12,11^. Thus, retinal WLR is not merely as a descriptive measure of local retinal vessel geometry but a clinically relevant biomarker that integrates cumulative cardiovascular risk.

Eutrophic vascular remodeling develops as an adaptive response to elevated blood pressure and increased myogenic vasoconstriction. In this case, the lumen of the arteriole narrows without an actual increase in the thickness of the vascular wall. Consequently, the WLR increases, while the wall cross-sectional area remains unchanged. As changes in WLR can be detected prior to wall hypertrophy, it serves as a sensitive early marker of retinal microvascular alterations ^32,12^.

In their study on factors influencing WLR in healthy, non-pregnant probands, Arichika et al. determined a mean WLR of 0.28 µm in their sample (> 50 years) ^32^. In our control group, the WLR was 0.19 µm (mean age 26 years). A robust correlation between WLR and age ^32,12^ has been demonstrated, which is in alignment with the findings of this study and provides a rationale for the observed discrepancy in WLR when comparing healthy pregnant women to our non-pregnant control group. In addition, a robust correlation between the WLR and both systolic and diastolic blood pressure has been demonstrated in the literature ^12,32^, which is consistent with the findings presented here for the correlation of WLR with MAP, which incorporates systolic and diastolic values in its calculation. The maternal cardiovascular system undergoes significant adaptation processes during pregnancy, characterized by increased blood volume, cardiac output, and MAP, especially in the third trimester ^35^. The continuous increase in WLR in the third trimester is most likely due to these adaptation processes and thus demonstrates that WLR is a sensitive marker to reflect the “natural stress test” for the maternal cardiovascular system during pregnancy. In contrast to the literature, no statistically significant relationship was observed between BMI and WLR in the present study ^12^. This discrepancy may be attributed to the fact that a significant proportion of the subjects had a normal BMI.

Changes during pregnancy differ between singleton and twin pregnancies primarily due to the greater physiological and hemodynamic demands present in twin gestations ^36–38^. However, the basic adaptive mechanisms remain similar, involving baseline blood pressure, vascular autoregulatory responses and thus microvascular remodeling to accommodate increased maternal and fetal needs ^39,40^. This may explain why the increase in WLR did not differ significantly between singletons and twin pregnancies.

Lupton et al. could not demonstrate any difference in retinal arteriolar caliber between nulliparous and multiparous women in their study using nonmydriatic fundus camera to measure total diameter of arteries ^41^. Our analysis revealed that primiparous individuals had a significantly lower WLR at the beginning of their pregnancy, with a steeper increase in WLR throughout gestation. Microvascular changes during pregnancy are influenced by parity, resulting in enhanced vascular remodeling, improved endothelial function and regulation of peripheral resistance and blood pressure, and altered autonomic regulation in multiparous women ^42^. In addition, parity is associated with a lower risk of pregnancy complications such as preeclampsia, linked to improved vascular adaptations affecting microvascular health ^42,43^. This could explain why primiparous women showed a significantly lower baseline WLR compared to nulliparous women. While primiparous women had a similar baseline WLR (0.186) to multiparous women (0.188), both values were significantly lower than those of nulliparous women (0.201), though the difference for multiparous women lacked significance, likely due to a smaller sample size (N=68). The results of the parity comparison suggest that, following one delivery before, physiological vascular remodeling processes have occurred to such an extent that baseline WLR values are lower at the start of subsequent pregnancies. In addition, nulliparous women appear to have a heightened response to MAP changes during pregnancy, resulting in more notable vascular remodeling. After at least one delivery, vascular adaptations remain responsive to MAP changes, though to a lesser degree due to established improved endothelial function and autonomic regulation.

Unfortunately, other studies on parity and vascular adaptation only differentiate between nulliparous and parous (≥1), without providing further details ^42,44^. Consequently, no comparison can be made with the differences between primiparous and multiparous women (≥ 2).

### Limitations

The most important limitation of the study is that no baseline WLR values were available prior to the start of pregnancy. Thus, the intercepts (0 weeks of gestation) had to be estimated using data mainly from the second and third trimester. Secondly, certain subgroups, e.g. twin pregnancies, and measurements in the first trimester, exhibited low case numbers. Thirdly, of the pregnant women in the first trimester who primarily attended our clinic for first trimester screening, the majority gave birth in other hospitals and did not return for further appointments during their pregnancy. Consequently, only a small number of cases of women with measurement intervals above more than 16 weeks could be included in the analysis of the intra-individual development of WLR during pregnancy.

### Perspectives

Adaptive optics–based measurement of retinal WLR provides a sensitive and reproducible means to quantify microvascular remodeling in vivo. For the first time, longitudinal changes in retinal microvasculature were investigated across trimesters in a well-phenotyped healthy cohort of pregnant women, and mean values of the WLR were defined for different gestational ages. There is a continuous increase in WLR over gestational age, which correlates statistically significantly with the MAP. The changes in retinal microvasculature during pregnancy should be interpreted not only as physiological adaptations but also as a dynamic stress test of the maternal vascular system.

Modifications in WLR have the promising potential to predict pregnancy-associated changes in an early phase, which is why the definition of mean values of WLR at different gestational times as a basis for comparison is of utmost relevance. Further longitudinal studies are required to investigate changes in WLR during pregnancy in risk groups, such as those with gestational diabetes or hypertensive pregnancy disorders, as well as in the postpartum period following pregnancy-associated complications, in order to predict individual cardiovascular disease risk. The follow-up of postpartum women following a healthy pregnancy may also facilitate a more nuanced understanding of the proportion of WLR that reflects reversible (functional) versus irreversible (structural) vascular remodeling.

## Novelty and Relevance

### What Is New?

‒ retinal microvasculature during pregnancy with cellular-level resolution was measured in-vivo by Adaptive Optics retinal Imaging

‒ longitudinal significant changes in Wall-to-Lumen ratio (WLR) in arterioles were demonstrated in healthy pregnant women with increasing gestational age

### What Is Relevant?

‒ continuous rise in WLR correlates significant with the mean arterial blood pressure

‒ WLR serves as indication of vascular stress test during pregnancy

### Clinical/Pathophysiological Implications?

‒ WLR provides as sensitive marker to quantify microvascular remodeling

‒ definition of normative data in healthy pregnant women allow distinction to microvasculature changes in pregnancy-associated complications and could predict individual cardiovascular risk postpartum

## Sources of Funding

This work was funded by the German Research Foundation (Deutsche Forschungsgemeinschaft, DFG); project number 493646873 – MD-LEICS. FG Rauscher is funded by the German Research Foundation (Deutsche Forschungsgemeinschaft, DFG); project number 497989466.

## Disclosures

The authors declare no conflict of interest.

## Data Availability

The data that support the findings of this study are not openly available due to reasons of sensitivity and are available from the corresponding author upon reasonable request. Data are located in controlled access data storage at Leipzig University, Medical Faculty.

## Notes

### Competing Interest Statement

The authors have declared no competing interest.

### Clinical Trial

German Clinical Trial Register number: DRKS00032530

### Author Declarations

Written informed consent was obtained from all participants. Research related to human use has been complied with all the relevant national regulations, institutional policies and in accordance the tenets of the Helsinki Declaration, and has been approved by Leipzig University?s institutional review board (IRB00001750, 052/23-ek).

## References

1. Facca TA, Mastroianni-Kirsztajn G, Sabino ARP, Passos MT, Dos Santos LF, Famá EAB, et al. Pregnancy as an early stress test for cardiovascular and kidney disease diagnosis. Pregnancy hypertension. 2018;12:169–173.

2. Osol G, Mandala M. Maternal uterine vascular remodeling during pregnancy. Physiology (Bethesda, Md.). 2009;24:58–71.

3. Osol G, Moore LG. Maternal uterine vascular remodeling during pregnancy. Microcirculation (New York, N.Y. : 1994). 2014;21:38–47.

4. Vargas AI, Tarraf SA, Jennings T, Bellini C, Amini R. Vascular Remodeling During Late-Gestation Pregnancy. An In-Vitro Assessment of the Murine Ascending Thoracic Aorta. Journal of biomechanical engineering. 2024;146.

5. Enkhmaa D, Wall D, Mehta PK, Stuart JJ, Rich-Edwards JW, Merz CNB, et al. Preeclampsia and Vascular Function. A Window to Future Cardiovascular Disease Risk. Journal of women’s health (2002). 2016;25:284–291.

6. Gifford FJ, Moroni F, Farrah TE, Hetherington K, MacGillivray TJ, Hayes PC, et al. The Eye as a Non-Invasive Window to the Microcirculation in Liver Cirrhosis. A Prospective Pilot Study. Journal of clinical medicine. 2020;9.

7. Kellner RL, Harris A, Ciulla L, Guidoboni G, Verticchio Vercellin A, Oddone F, et al. The Eye as the Window to the Heart. Optical Coherence Tomography Angiography Biomarkers as Indicators of Cardiovascular Disease. Journal of clinical medicine. 2024;13.

8. Zhang J, Shi L, Shen Y. The retina. A window in which to view the pathogenesis of Alzheimer’s disease. Ageing research reviews. 2022;77:101590.

9. Burns SA, Elsner AE, Sapoznik KA, Warner RL, Gast TJ. Adaptive optics imaging of the human retina. Progress in retinal and eye research. 2019;68:1–30.

10. Bedggood P, Metha A. Adaptive optics imaging of the retinal microvasculature. Clinical & experimental optometry. 2020;103:112–122.

11. Ciuceis C de, Rosei CA, Malerba P, Rossini C, Nardin M, Chiarini G, et al. Prognostic significance of the wall to lumen ratio of retinal arterioles evaluated by adaptive optics. European journal of internal medicine. 2024;122:86–92.

12. Meixner E, Michelson G. Measurement of retinal wall-to-lumen ratio by adaptive optics retinal camera. A clinical research. Graefe’s archive for clinical and experimental ophthalmology = Albrecht von Graefes Archiv fur klinische und experimentelle Ophthalmologie. 2015;253:1985–1995.

13. Pinhas A, Dubow M, Shah N, Chui TY, Scoles D, Sulai YN, et al. In vivo imaging of human retinal microvasculature using adaptive optics scanning light ophthalmoscope fluorescein angiography. Biomedical optics express. 2013;4:1305–1317.

14. Dathan-Stumpf A, Stepan H, Valterova E, Jakubicek R, Berbée C, Seidenspinner M-L, et al. Pregnancy induces retinal microvascular changes indicating cardio-metabolic stress. Pregnancy hypertension. 2024;35:30–31.

15. Wang X, Wang X, Chou Y, Ma J, Zhong Y. Significant retinal microvascular impairments in multiple sclerosis assessed through optical coherence tomography angiography. Multiple sclerosis and related disorders. 2023;70:104505.

16. Assialioui A, Marco-Pascual C, Torrente-Segarra V, Domínguez R, Santos N, Peñafiel J, et al. Microvascular abnormalities in skin capillaries of individuals with amyotrophic lateral sclerosis. Scientific reports. 2024;14:24648.

17. Arbeitsgemeinschaft Geburtshilfe und Pränatalmedizin in der DGGGG. S3-Leitlinie Gestationsdiabetes mellitus (GDM), Diagnostik, Therapie und Nachsorge. AWMF-Registernummer: 057–008. 2019:https://register.awmf.org/assets/guidelines/057-008l_S3_Gestationsdiabetes-mellitus-GDM-Diagnostik-Therapie-Nachsorge_2019-06.pdf.

18. Pagsibigan JS, Balabagno AO, Tuazon JA, Evangelista LS. Blood Pressure Measurement Training Program and Adherence of Public Health Nurses to BP Measurement Guidelines. Acta medica Philippina. 2017;51:351–359.

19. Williams B, Mancia G, Spiering W, Agabiti Rosei E, Azizi M, Burnier M, et al. 2018 ESC/ESH Guidelines for the management of arterial hypertension. European heart journal. 2018;39:3021–3104.

20. National Institute for Health and Care Excellence (NICE). Hypertension in adults: diagnosis and management. 2023:www.nice.org.uk/guidance/ng136, aufgerufen 25.06.2024.

21. S3-Leitlinie: Nationale Versorgungsleitlinie Hypertonie. Registernummer nvl - 009. 2023:https://register.awmf.org/de/leitlinien/detail/nvl-009, aufgerufen 25.06.2024.

22. Weichert A, Hagen A, Tchirikov M, Fuchs IB, Henrich W, Entezami M. Reference Curve for the Mean Uterine Artery Pulsatility Index in Singleton Pregnancies. Geburtshilfe und Frauenheilkunde. 2017;77:516–523.

23. Cavoretto PI, Salmeri N, Candiani M, Farina A. Reference ranges of uterine artery pulsatility index from first to third trimester based on serial Doppler measurements. Longitudinal cohort study. Ultrasound in obstetrics & gynecology : the official journal of the International Society of Ultrasound in Obstetrics and Gynecology. 2023;61:474–480.

24. Cristescu IE, Zagrean L, Balta F, Branisteanu DC. RETINAL MICROCIRCULATION INVESTIGATION IN TYPE I AND II DIABETIC PATIENTS WITHOUT RETINOPATHY USING AN ADAPTIVE OPTICS RETINAL CAMERA. Acta endocrinologica (Bucharest, Romania : 2005). 2019;15:417–422.

25. Zaleska-Żmijewska A, Piątkiewicz P, Śmigielska B, Sokołowska-Oracz A, Wawrzyniak ZM, Romaniuk D, et al. Retinal Photoreceptors and Microvascular Changes in Prediabetes Measured with Adaptive Optics (rtx1™). A Case-Control Study. Journal of diabetes research. 2017;2017:4174292.

26. Baniasadi N, Rauscher FG, Li D, Wang M, Choi EY, Wang H, et al. Norms of Interocular Circumpapillary Retinal Nerve Fiber Layer Thickness Differences at 768 Retinal Locations. Translational vision science & technology. 2020;9:23.

27. Wang M, Elze T, Li D, Baniasadi N, Wirkner K, Kirsten T, et al. Age, ocular magnification, and circumpapillary retinal nerve fiber layer thickness. Journal of biomedical optics. 2017;22:1–19.

28. Scheibe P, Zocher MT, Francke M, Rauscher FG. Analysis of foveal characteristics and their asymmetries in the normal population. Experimental eye research. 2016;148:1–11.

29. Kupis M, Wawrzyniak ZM, Szaflik JP, Zaleska-Żmijewska A. Retinal Photoreceptors and Microvascular Changes in the Assessment of Diabetic Retinopathy Progression. A Two-Year Follow-Up Study. Diagnostics (Basel, Switzerland). 2023;13.

30. Bakker E, Dikland FA, van Bakel R, Andrade De Jesus D, Sánchez Brea L, Klein S, et al. Adaptive optics ophthalmoscopy. A systematic review of vascular biomarkers. Survey of ophthalmology. 2022;67:369–387.

31. Ciuceis C de, Coschignano MA, Caletti S, Rossini C, Duse S, Docchio F, et al. [PP.09.17] Reproducibility of the evaluation of the wall to lumen ratio of retinal arterioles with two different non-invasive approaches. Journal of Hypertension. 2017;35:e153–e154.

32. Arichika S, Uji A, Ooto S, Muraoka Y, Yoshimura N. Effects of age and blood pressure on the retinal arterial wall, analyzed using adaptive optics scanning laser ophthalmoscopy. Scientific reports. 2015;5:12283.

33. Isensee F, Jaeger PF, Kohl SAA, Petersen J, Maier-Hein KH. nnU-Net. A self-configuring method for deep learning-based biomedical image segmentation. Nature methods. 2021;18:203–211.

34. Jakubicek R, Rauscher F G, Odstrcilik J, Kolar R. An automated approach for computing the wall-to-lumen ratio from adaptive optics retinal images:in submission.

35. Mehta LS, Warnes CA, Bradley E, Burton T, Economy K, Mehran R, et al. Cardiovascular Considerations in Caring for Pregnant Patients. A Scientific Statement From the American Heart Association. Circulation. 2020;141:e884–e903.

36. Choi BY, Jung SY, Lee HK, Lee MJ, Kim HJ, Park JY, et al. Comparison of body composition change, measured with bioelectrical impedance analysis, between singleton and twin pregnancy. A prospective cohort study. European journal of obstetrics, gynecology, and reproductive biology. 2025;306:154–159.

37. Giorgione V, Melchiorre K, O’Driscoll J, Khalil A, Sharma R, Thilaganathan B. Maternal echocardiographic changes in twin pregnancies with and without pre-eclampsia. Ultrasound in obstetrics & gynecology : the official journal of the International Society of Ultrasound in Obstetrics and Gynecology. 2022;59:619–626.

38. Umazume T, Yamada T, Furuta I, Iwano H, Morikawa M, Watari H, et al. Morphofunctional cardiac changes in singleton and twin pregnancies. A longitudinal cohort study. BMC pregnancy and childbirth. 2020;20:750.

39. Sima R-M, Findeklee S, Bădărău I-A, Poenaru M-O, Scheau C, Pleș L. Comparison of maternal third trimester hemodynamics between singleton pregnancy and twin pregnancy. Journal of perinatal medicine. 2021;49:566–571.

40. Meah VL, Kimber ML, Khurana R, Howse R, Hornberger LK, Steinback CD, et al. Cardioautonomic control in healthy singleton and twin pregnancies. *Journal of applied physiology* (Bethesda, Md. : 1985). 2021;130:923–932.

41. Lupton SJ, Chiu CL, Hodgson LAB, Tooher J, Lujic S, Ogle R, et al. Temporal changes in retinal microvascular caliber and blood pressure during pregnancy. Hypertension (Dallas, Tex. : 1979). 2013;61:880–885.

42. Morris EA, Hale SA, Badger GJ, Magness RR, Bernstein IM. Pregnancy induces persistent changes in vascular compliance in primiparous women. American journal of obstetrics and gynecology. 2015;212:633.e1–6.

43. Ling HZ, Guy GP, Bisquera A, Poon LC, Nicolaides KH, Kametas NA. The effect of parity on longitudinal maternal hemodynamics. American journal of obstetrics and gynecology. 2019;221:249.e1–249.e14.

44. Iizuka M, Miyasaka N, Hirose Y, Toba M, Sakamoto S, Kubota T. Is there a differential impact of parity on factors regulating maternal peripheral resistance? Hypertension research : official journal of the Japanese Society of Hypertension. 2016;39:737–743.

